# Links of Common Infections with Sleep in Middle-Aged and Older Adults: Modification by Race in the Baltimore Epidemiologic Catchment Area Study

**DOI:** 10.64898/2026.01.14.26344029

**Authors:** Yiwei Yue, Jill A. Rabinowitz, Chandra L. Jackson, Yiping Xia, Idiatou Diallo, Robert Yolken, William W. Eaton, Brion S. Maher, Adam P. Spira

**Affiliations:** Department of Mental Health, Johns Hopkins Bloomberg School of Public Health, Baltimore, MD USA; Department of Psychiatry, Robert Wood Johnson Medical School, Rutgers University, Piscataway, NJ, USA; Epidemiology Branch, National Institute of Environmental Health Sciences, National Institutes of Health, Department of Health and Human Services, Research Triangle Park, Durham, North Carolina, USA; Intramural Research Program, National Institute on Minority Health and Health Disparities, Bethesda, Maryland, USA; Stanley Laboratory of Developmental Neurovirology, Department of Pediatrics, Johns Hopkins School of Medicine, Baltimore, MD USA; Department of Psychiatry and Behavioral Sciences, Johns Hopkins University School of Medicine, Baltimore, MD USA; Center on Aging and Health, Johns Hopkins University, Baltimore, MD USA

**Keywords:** Toxoplasma gondii, adults, racial disparities, infectious diseases, herpesviruses, health, cohort studies, minority groups

## Abstract

**Objectives:** To determine associations of common infections with self-reported sleep and explore whether these links differ by race in a diverse cohort.

**Methods:** We studied 602 participants from the Baltimore Epidemiologic Catchment Area Study (mean age 59.0±12.8 years; 36.9% male; 35.6% minoritized adults) with data on common infections (IgG antibodies to herpes simplex virus type 1, cytomegalovirus, varicella zoster virus, Epstein-Barr virus, and *Toxoplasma gondii*) and self-reported symptoms of insomnia (i.e., difficulty falling or staying asleep and early awakening), hypersomnia, and sleep duration.

**Results:** We observed differences by race in the association of common infections with sleep disturbances (p-values for interactions<0.05). In fully-adjusted models, among White participants, those who were seropositive for CMV had 51% higher odds of insomnia symptoms (OR=1.51, 95% CI: 0.88, 2.60) and 26% higher odds of early awakening (OR=1.26, 95% CI: 0.67, 2.39). Among minoritized adults, however, CMV seropositivity was associated with 75% lower odds of insomnia symptoms (OR=0.25, 95% CI: 0.09, 0.67) and 73% lower odds of early awakening (OR=0.27, 95% CI: 0.10, 0.74). Seropositivity for a higher number of infections (β=0.24, 95% CI: -0.01, 0.48) or for TOX alone (β=0.64, 95% CI: 0.08, 1.20) was associated with longer sleep among minoritized participants, but shorter sleep among White participants.

**Conclusions:** In middle-aged and older adults, common infections are differentially associated with sleep disturbances and duration, with infections linked to better sleep profiles and longer sleep among minoritized adults, but poorer sleep among White adults. Research is needed to identify the sources of these differences.

## Introduction

Sleep disturbances are common in later life^1^ and are associated with increased risk for adverse health outcomes, including mental and physical health conditions (e.g., depression, obesity, diabetes, cardiovascular disease) and mortality.^2,3^ Compared to younger populations, middle-aged and older adults exhibit more sleep disturbances such as shorter sleep duration, lower sleep efficiency, and more wakefulness after sleep onset.^4–6^ Given the numerous sleep-related health sequalae among middle-aged and older adults, identifying factors that contribute to sleep disturbances could inform interventions to improve sleep health in this population.

Infections, including herpesviruses (i.e., herpes simplex virus type 1 (HSV-1), cytomegalovirus (CMV), Epstein-Barr virus (EBV), varicella zoster virus (VZV)) and *Toxoplasma gondii* (TOX), are common, and become more prevalent as individuals age.^7–11^ For example, a study found that 45.2% of middle-aged adults (39-45 years) and 56.5% of older adults (64-70 years) are seropositive for CMV.^12^ In the same study, seropositivity rates for EBV were 81.7% and 76.1% for middle-aged and older adults, respectively.^12^ Other studies have shown that approximately 90% of older adults show serological evidence of HSV-1, CMV, and EBV,^13^ and TOX, a potentially neurotropic intracellular protozoan, has been observed in approximately 25% of the U.S. population.^10^ Numerous studies have linked common infections to earlier onset and severity of psychiatric conditions^14–21^; however, less is known about the association of common infections with sleep disturbances among middle-aged and older adults.

There are a number of reasons to expect that common infections may be associated with sleep disturbances. Common infections may contribute to chronic and systemic inflammation, linked to increased inflammatory cytokine levels, and influence neurotransmitter systems (e.g., dopamine) that regulate sleep-wake cycles.^22,23,24^ Moreover, common infections have been associated with decrements in depressed mood and fatigue, known risk factors for insomnia and sleep disturbances.^24,25^ In addition, sleep disturbances can increase the risk of common infections. Sleep deprivation or insufficient sleep has been causally linked to greater risk of infection, as it makes individuals more vulnerable to infection.^26–29^ Further, short sleep duration has been linked to greater proinflammatory cytokines, which may contribute to greater susceptibility to acquiring infections.^30,31^ Thus, it is likely that the relationship between common infections and sleep is bi-directional.

Prior studies examining associations of common infections with sleep disturbances have primarily focused on the association of TOX with sleep and have yielded mixed results. For example, in a clinical sample, adults aged 18 to 80 years (mean = 43.63 years) with insomnia had higher levels of TOX antibodies compared to individuals without insomnia.^23^ In contrast, another study reported that adults seropositive for TOX endorsed fewer sleep problems, and that higher TOX titers were associated with longer sleep duration.^32^ Other studies have found no differences in insomnia symptoms, sleep onset latency, or sleep duration by TOX seropositivity.^33,34^ Research has also shown that the prevalence of herpes zoster was higher among adults with sleep disorders, and that those with sleep disorders had a higher risk of incident herpes zoster compared to those without.^7^ Discrepancies in study findings may be due in part to variation in the demographic characteristics of their samples, including age and race. Regardless, the minimal research on infection and sleep among middle-aged and older adults and inconsistencies across study findings highlight the need for further research.

Moreover, few studies have examined whether demographic factors, such as race, influence associations of common infections with sleep disturbances among older adults. Compared to White populations, historically and persistently marginalized groups, including African Americans, report poorer sleep quality,^35,36^ shorter sleep duration,^35–41^ more sleep fragmentation,^35^ and lower sleep efficiency.^39^ In addition, relative to White adults, minoritized adults have higher CMV antibody levels and a higher incidence of CMV seropositivity,^8,42^ an increased odds of HSV-1 seropositivity,^43^ and higher infection burden (i.e., a higher number of different infections).^44^ These disparities may be due, at least in part, to economic disadvantage and structural barriers (e.g., crowded living conditions) that exacerbate risk for both infection transmission and sleep disturbance.

In the current study, we investigated associations of common infections with sleep characteristics in a diverse, population-based cohort of older adults. Compared to younger populations, middle-aged and older adults tend to experience greater sleep disturbance and are more vulnerable to acquiring infections,^1–11^ highlighting the importance of further understanding the associations of common infections with sleep disturbances in middle-aged and older people. We also determined whether race moderated links between common infections and sleep disturbances to examine whether infections are potential drivers of disparities in sleep health.

## Methods

### Participants

We studied participants from the Epidemiologic Catchment Area (ECA) study, a population-based, longitudinal study launched in 1981 to identify the prevalence and incidence of psychiatric disorders in five major cities in the US, including Baltimore, Maryland; New Haven, Connecticut, St. Louis, Missouri; Durham, North Carolina; and Los Angeles, California.^45^ Data from the current study were drawn from the Baltimore site. Five waves of data collection occurred at the Baltimore site, starting in 1981 (Wave 1), 1982 (Wave 2), 1993-1996 (Wave 3), 2003-2006 (Wave 4), and 2017-2022 (Wave 5).

To be eligible to enroll in the study in 1981, participants had to be ≥ 18 years old. We restricted the current analyses to 602 participants from Wave 4 who had valid data from serum antibodies and sleep measures (see below). The Baltimore ECA study adhered to the Declaration of Helsinki principles and was approved by the Institutional Review Board of the Johns Hopkins Bloomberg School of Public Health. Written informed consent was obtained from all participants.

### Collection of biological samples, antibody levels, and infection seropositivity

At Wave 4, 888 (approximately 83%) of Baltimore ECA participants (N=1,071) agreed to donate a biological sample in blood or buccal specimen form, and 683 participants consented to donate a blood sample.^46^ Immunoglobin G (IgG) antibodies to HSV-1, CMV, EBV, VZV, and TOX were measured using solid phase immunoassays as previously described.^47,48^ To identify antibody levels and seropositivity, we used the manufacturer’s designated thresholds to determine whether participants were exposed to the corresponding infectious agents.^48^ The criteria employed to determine higher serum IgG levels involved examining whether the distribution of IgG levels was in the highest quintile, based on thresholds determined by IgG distributions of control participants. In addition to separate variables reflecting seropositivity for each of the infections, we created a variable reflecting the sum of the total number of positive antibody tests.

### Sleep characteristics

Symptoms of insomnia, early awakening, and hypersomnia, were assessed using items from the depression module of the Diagnostic Interview Schedule (DIS), a lay-interviewer-administered structured interview for Diagnostic and Statistical Manual of Mental Disorders, Revised Third Edition (DSM-III-R) diagnoses.^49^ Insomnia was assessed via the following question: “Have you ever had two weeks or more when nearly every night you had trouble falling asleep, staying asleep, or waking up too early?” In addition to appearing in the insomnia item, early awakening was measured separately from other insomnia symptoms: “Have you ever had two weeks or more when nearly every morning, you would wake up at least 2 hours before you wanted to?” We chose to analyze this item as an outcome, in addition to the other insomnia symptom item, because it could help determine the extent to which infection is associated with insomnia more broadly, or if an infection-insomnia association is driven by early awakening. Hypersomnia was also assessed: “Have you ever had two weeks or longer when nearly every day you were sleeping too much?” The response options for these items were yes/no. Each item was coded as a binary variable (0 = symptom absence, 1 = presence). Participants also were asked how many hours of sleep they usually obtained in a 24-hour period. Answers were given in integers, and we used this as a continuous measure of 24-hour sleep duration.

### Potential modifier: Race/ethnicity

Race was measured based on self-identification as American Indian, Alaskan Native, Asian, Pacific Islander, Black/not Hispanic, Hispanic, or White/not Hispanic.

### Sociodemographic and Clinical Characteristics

Interviewers recorded additional participant sociodemographic characteristics including age, sex (as observed), years of educational attainment (ranging from 0 (none) to 17 (graduate school)), annual household income (0 = no income; 1 = less than $1,000; 2 = $1,000-$1,999 … 12 = $11,000-$12,499; 13 = $12,500-$14,999; 14 = $15,000-$17,499; 15 = $17,500-$19,999; 16 = $20,000-$24,999; 17 = $25,000-$34,999, 18 = $35,000-$49,999; 19 = $50,000-$64,999; 20 = $65,000-$79,999; 21 = $80,000-$89,999; 22 = $90,000-$99,999; 23 = $100,000-$124,999; 24 = $125,000-$149,999; 25 = $150,000-$199,999; 26 = $200,000 and over), and household size (total number of people lived in the participant’s household). In terms of clinical characteristics, participants were asked about lifetime comorbidities including high blood pressure, heart disease (i.e., rheumatic heart disease, angina pectoris, congestive heart failure), high blood sugar or diabetes, and smoking status. BMI was calculated using objective measurements of participants’ weight and height. However, some participants refused to have their weight or height measured or their weight exceeded the limits on the scale. For these participants, self-reported weight and height were used to determine their BMI.

### Statistical Analyses

We calculated descriptive statistics and examined whether racial differences existed in seropositivity for infection and sleep using Pearson’s chi-squared tests and *t*-tests. We created a binary variable reflecting individuals who identified as White (0) or who identified as American Indian, Alaskan Native, Asian, Pacific Islander, Black- not Hispanic, or Hispanic (1 = minoritized population).

To examine the association of seropositivity for common infections (i.e., HSV-1, CMV, VZV, EBV, TOX, infection count) with binary sleep disturbance variables (i.e., insomnia symptoms, early awakening, hypersomnia), we conducted logistic regression analyses; we performed linear regression analyses to examine associations of infection with sleep duration. Minimally adjusted models included age, sex, race, and years of education (Model 1). Fully adjusted models adjusted further for BMI, diabetes, hypertension, heart problems, household income, household size, and smoking status (Model 2). Three binary variables reflecting the presence or absence of diabetes, hypertension, and heart problems, (0 = absence of condition, 1 = presence). We also created a categorical smoking status variable (non-smoker = 0, former smoker (≥ 6 months since tobacco use) = 1, current smoker = 2). All continuous variables including age, education, BMI, household income and size were centered prior to model fitting.

To determine whether race moderated associations between common infections and various sleep outcomes, we included two-way interaction terms (infection X race) in minimally- and fully-adjusted models. We only report interaction results when infection X race interaction terms were statistically significant (p < 0.05) and obtained model-derived point estimates to determine the direction and magnitude of associations between infections and outcomes by race. All analyses were conducted using Stata software version SP 17.0 (StataCorp, College Station, TX).

## Results

Participants (N = 602) were 59.0 ± 12.8 years of age (range: 41-97), 36.9% (N = 222) were male, and 35.6% (N = 214) were minoritized adults who identified as either American Indian (n = 10, 4.7%), Asian (n = 7, 3.3%), Pacific Islander (n = 2, 0.9%), Black not Hispanic (n = 190, 88.8%), or Hispanic (n = 5, 2.3%) (Table 1). Overall, 68.8% of participants were seropositive for HSV-1, 67.9% for CMV, 79.6% for EBV, 20.3% for VZV, and 24.6% for TOX. The mean number of positive antibody tests was 2.62 ± 1.07 (range = 0-5). A higher proportion of minoritized vs. White adults were seropositive for CMV (83.64% vs. 59.28%; χ^2^(1) = 37.5966, p < 0.001) and EBV (88.32% vs. 74.74%; χ^2^(1) = 15.6355, p < 0.001), and minoritized participants had a higher total number of positive antibody tests vs. White participants (2.86 vs. 2.48; *t* = -4.1495, p < 0.001). Minoritized participants were less likely than White participants to report insomnia symptoms (19.16% vs. 28.61%; χ^2^(2) = 6.5360, p = 0.038). Differences in key study variables by infection status was shown in Supplementary Table 1.

**Table 1.**
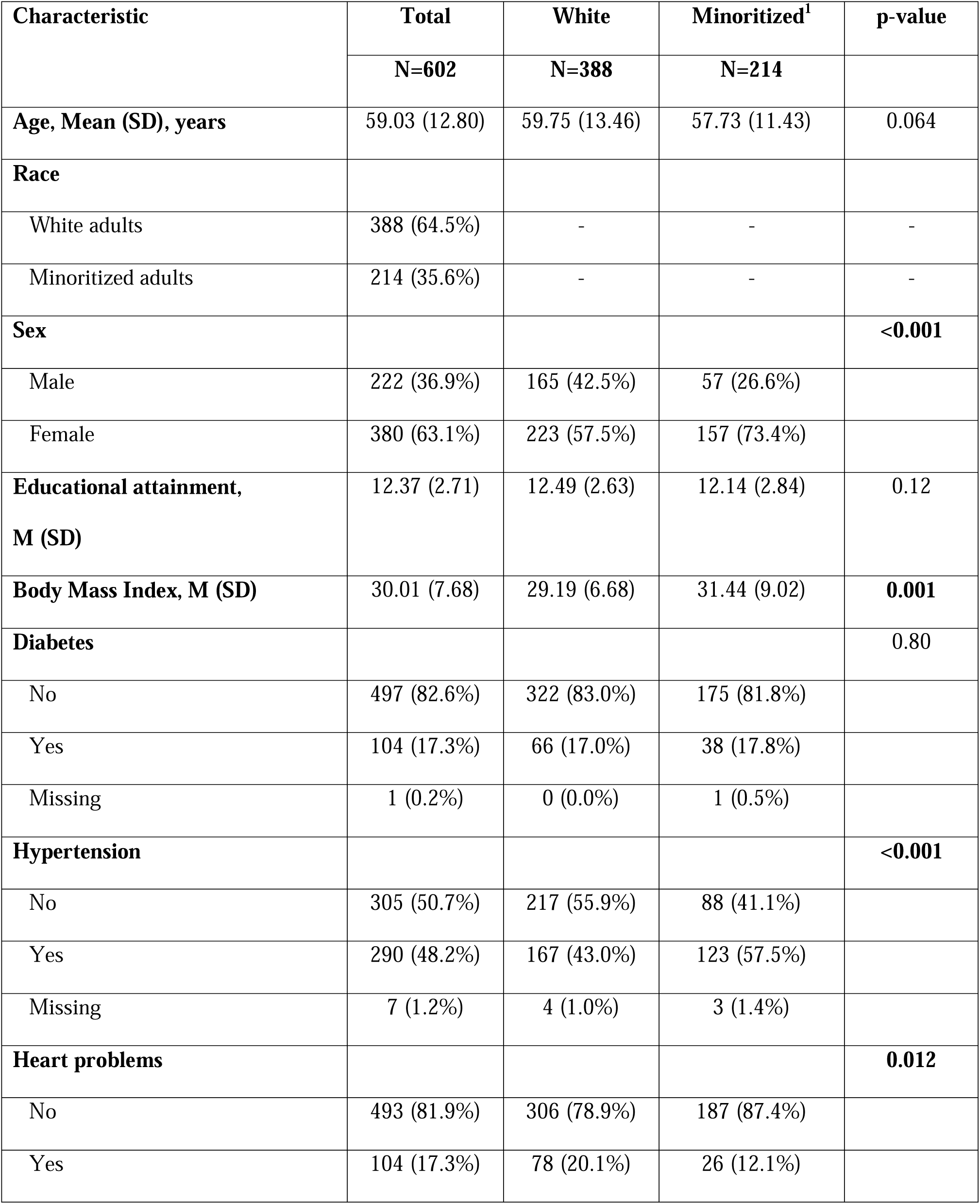
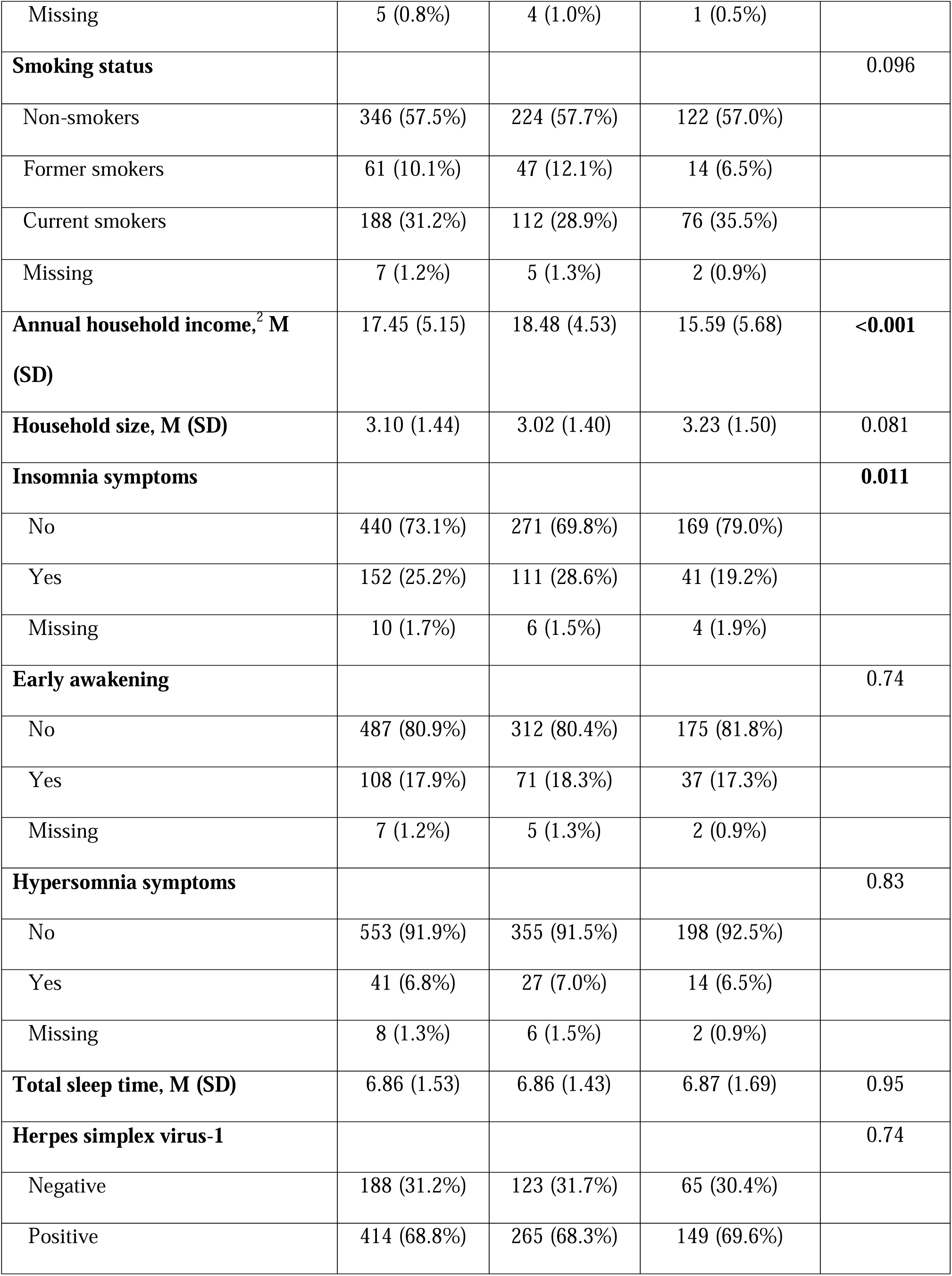
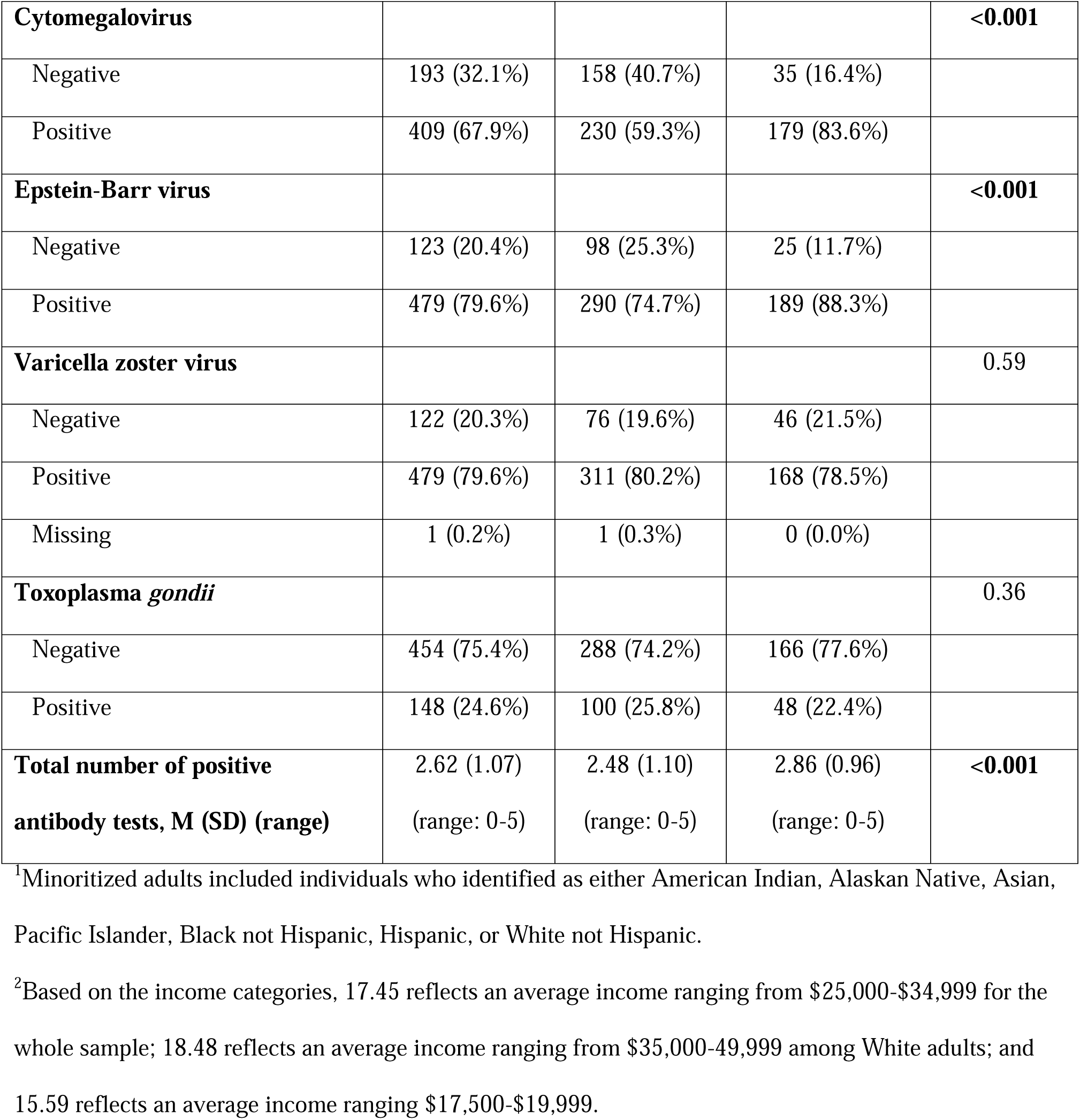
Study Population Characteristics Overall and by Race, Baltimore Epidemiologic Catchment Area (ECA) study Wave 4 (2003-2006)

| Characteristic | Total | White | Minoritized<sup>1</sup> | p-value |
| --- | --- | --- | --- | --- |
|  | N=602 | N=388 | N=214 |  |
| Age, Mean (SD), years | 59.03 (12.80) | 59.75 (13.46) | 57.73 (11.43) | 0.064 |
| **Race** |  |  |  |  |
| White adults | 388 (64.5%) | - | - | - |
| Minoritized adults | 214 (35.6%) | - | - | - |
| **Sex** |  |  |  | **<0.001** |
| Male | 222 (36.9%) | 165 (42.5%) | 57 (26.6%) |  |
| Female | 380 (63.1%) | 223 (57.5%) | 157 (73.4%) |  |
| **Educational attainment, M (SD)** | 12.37 (2.71) | 12.49 (2.63) | 12.14 (2.84) | 0.12 |
| **Body Mass Index, M (SD)** | 30.01 (7.68) | 29.19 (6.68) | 31.44 (9.02) | **0.001** |
| **Diabetes** |  |  |  | 0.80 |
| No | 497 (82.6%) | 322 (83.0%) | 175 (81.8%) |  |
| Yes | 104 (17.3%) | 66 (17.0%) | 38 (17.8%) |  |
| Missing | 1 (0.2%) | 0 (0.0%) | 1 (0.5%) |  |
| **Hypertension** |  |  |  | **<0.001** |
| No | 305 (50.7%) | 217 (55.9%) | 88 (41.1%) |  |
| Yes | 290 (48.2%) | 167 (43.0%) | 123 (57.5%) |  |
| Missing | 7 (1.2%) | 4 (1.0%) | 3 (1.4%) |  |
| **Heart problems** |  |  |  | **0.012** |
| No | 493 (81.9%) | 306 (78.9%) | 187 (87.4%) |  |
| Yes | 104 (17.3%) | 78 (20.1%) | 26 (12.1%) |  |
| Missing | 5 (0.8%) | 4 (1.0%) | 1 (0.5%) |  |
| **Smoking status** |  |  |  | 0.096 |
| Non-smokers | 346 (57.5%) | 224 (57.7%) | 122 (57.0%) |  |
| Former smokers | 61 (10.1%) | 47 (12.1%) | 14 (6.5%) |  |
| Current smokers | 188 (31.2%) | 112 (28.9%) | 76 (35.5%) |  |
| Missing | 7 (1.2%) | 5 (1.3%) | 2 (0.9%) |  |
| **Annual household income,<sup>2</sup> M (SD)** | 17.45 (5.15) | 18.48 (4.53) | 15.59 (5.68) | **<0.001** |
| **Household size, M (SD)** | 3.10 (1.44) | 3.02 (1.40) | 3.23 (1.50) | 0.081 |
| **Insomnia symptoms** |  |  |  | **0.011** |
| No | 440 (73.1%) | 271 (69.8%) | 169 (79.0%) |  |
| Yes | 152 (25.2%) | 111 (28.6%) | 41 (19.2%) |  |
| Missing | 10 (1.7%) | 6 (1.5%) | 4 (1.9%) |  |
| **Early awakening** |  |  |  | 0.74 |
| No | 487 (80.9%) | 312 (80.4%) | 175 (81.8%) |  |
| Yes | 108 (17.9%) | 71 (18.3%) | 37 (17.3%) |  |
| Missing | 7 (1.2%) | 5 (1.3%) | 2 (0.9%) |  |
| **Hypersomnia symptoms** |  |  |  | 0.83 |
| No | 553 (91.9%) | 355 (91.5%) | 198 (92.5%) |  |
| Yes | 41 (6.8%) | 27 (7.0%) | 14 (6.5%) |  |
| Missing | 8 (1.3%) | 6 (1.5%) | 2 (0.9%) |  |
| **Total sleep time, M (SD)** | 6.86 (1.53) | 6.86 (1.43) | 6.87 (1.69) | 0.95 |
| **Herpes simplex virus-1** |  |  |  | 0.74 |
| Negative | 188 (31.2%) | 123 (31.7%) | 65 (30.4%) |  |
| Positive | 414 (68.8%) | 265 (68.3%) | 149 (69.6%) |  |
| **Cytomegalovirus** |  |  |  | **<0.001** |
| Negative | 193 (32.1%) | 158 (40.7%) | 35 (16.4%) |  |
| Positive | 409 (67.9%) | 230 (59.3%) | 179 (83.6%) |  |
| **Epstein-Barr virus** |  |  |  | **<0.001** |
| Negative | 123 (20.4%) | 98 (25.3%) | 25 (11.7%) |  |
| Positive | 479 (79.6%) | 290 (74.7%) | 189 (88.3%) |  |
| **Varicella zoster virus** |  |  |  | 0.59 |
| Negative | 122 (20.3%) | 76 (19.6%) | 46 (21.5%) |  |
| Positive | 479 (79.6%) | 311 (80.2%) | 168 (78.5%) |  |
| Missing | 1 (0.2%) | 1 (0.3%) | 0 (0.0%) |  |
| **Toxoplasma gondii** |  |  |  | 0.36 |
| Negative | 454 (75.4%) | 288 (74.2%) | 166 (77.6%) |  |
| Positive | 148 (24.6%) | 100 (25.8%) | 48 (22.4%) |  |
| **Total number of positive antibody tests, M (SD) (range)** | 2.62 (1.07)<br>(range: 0-5) | 2.48 (1.10)<br>(range: 0-5) | 2.86 (0.96)<br>(range: 0-5) | **<0.001** |

<sup>1</sup>Minoritized adults included individuals who identified as either American Indian, Alaskan Native, Asian, Pacific Islander, Black not Hispanic, Hispanic, or White not Hispanic.
<sup>2</sup>Based on the income categories, 17.45 reflects an average income ranging from \$25,000-\$34,999 for the whole sample; 18.48 reflects an average income ranging from \$35,000-49,999 among White adults; and 15.59 reflects an average income ranging \$17,500-\$19,999.

Overall, there were no statistically significant associations of infections with any sleep outcomes in minimally or fully adjusted models (Table 2); however, we observed several interactions of infection with race (Table 3). Race moderated the association of CMV seropositivity with insomnia symptoms (interaction term p-value = 0.001) such that CMV seropositivity was associated with a 75% lower odds of insomnia symptoms (OR = 0.25, 95% CI: 0.09, 0.67, p-value = 0.006) among minoritized participants. Among White participants, this association was in the opposite direction; CMV was associated with a 51% higher odds of insomnia symptoms, although this association did not reach statistical significance (OR = 1.51, 95% CI: 0.88, 2.60, p-value = 0.138). We also observed an interaction between CMV seropositivity and race with regard to early awakening (interaction term p-value = 0.01). Among minoritized participants, CMV seropositivity was associated with a 73% lower odds of early awakening (OR = 0.27, 95% CI: 0.10, 0.74, p-value = 0.011); this association was in the opposite direction among White participants (OR = 1.26, 95% CI: 0.67, 2.39, p-value = 0.476). Although this association was not statistically significant, results suggest that White adults who were seropositive for CMV had a 26% higher odds of early awakening.

**Table 2.**
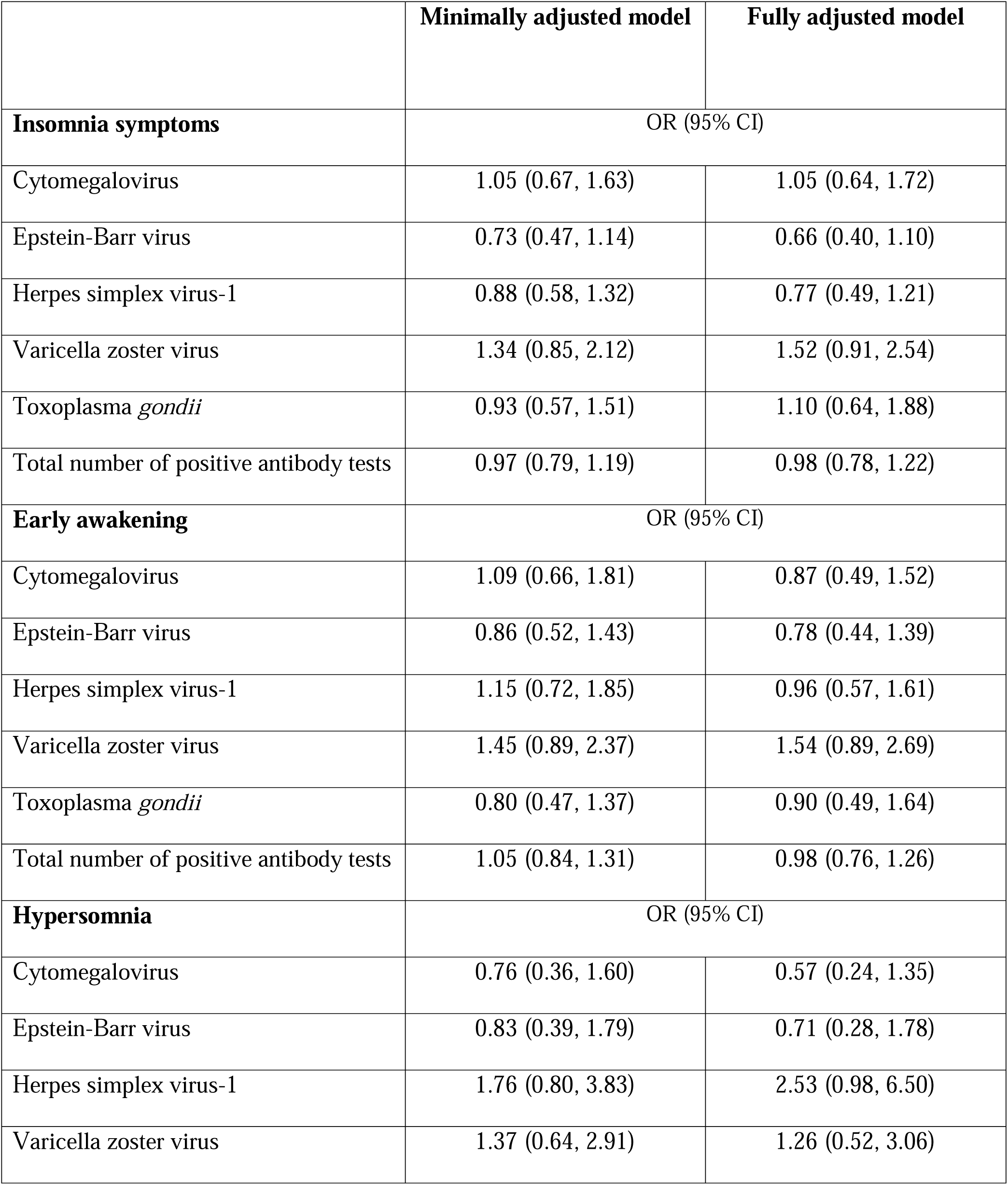
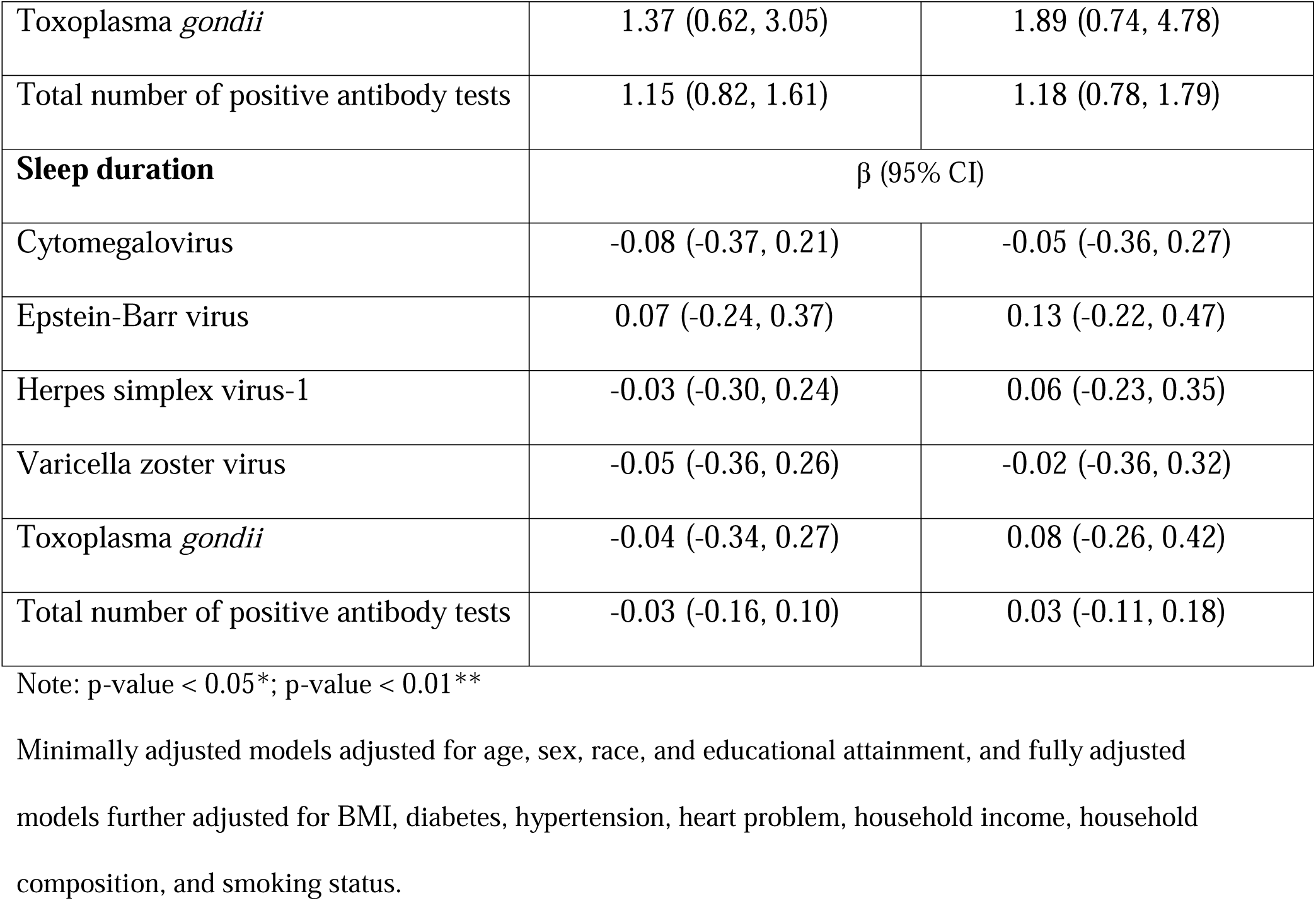
Associations between Common Infections and Sleep Disturbances.

**Table 3.**
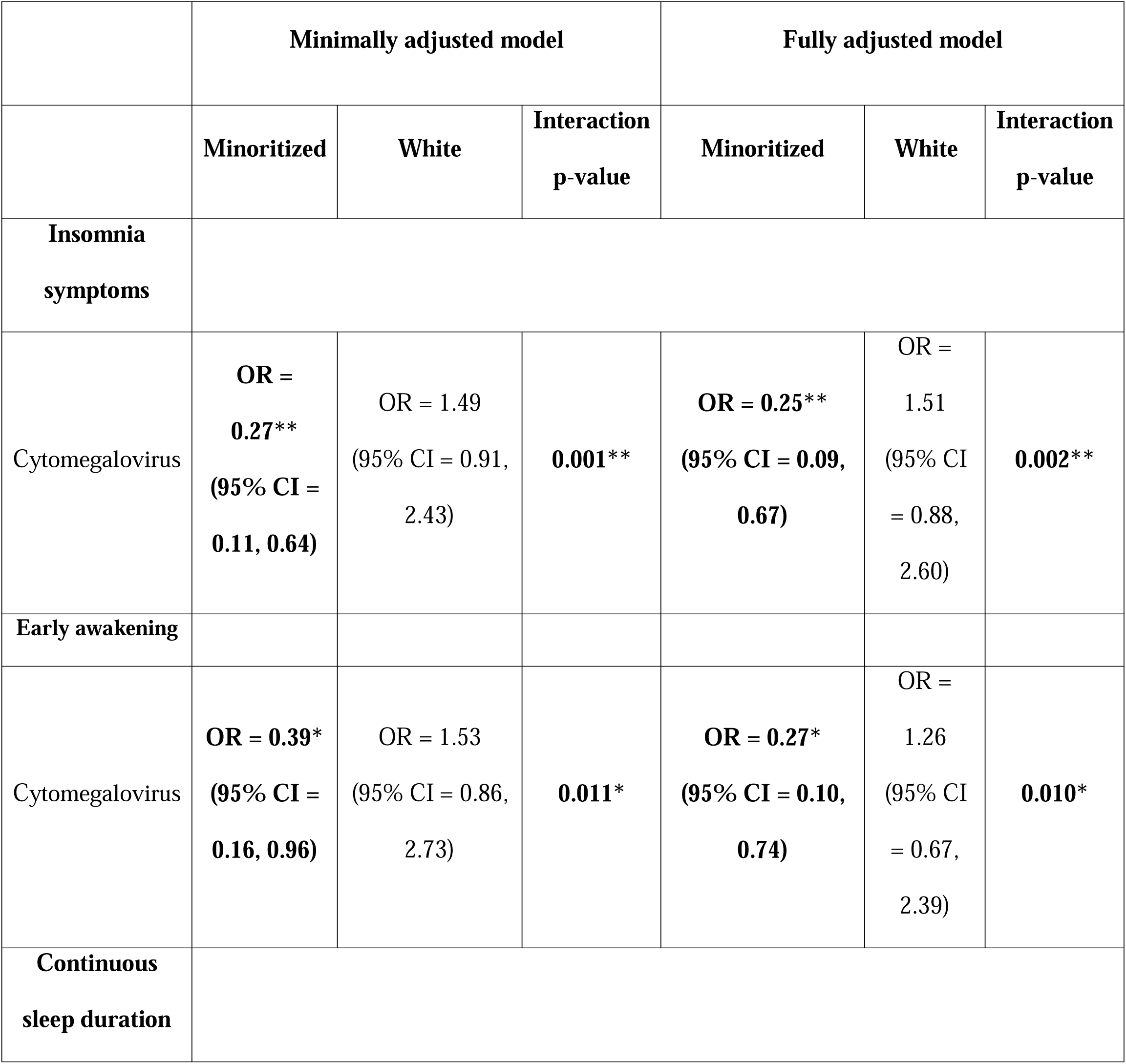
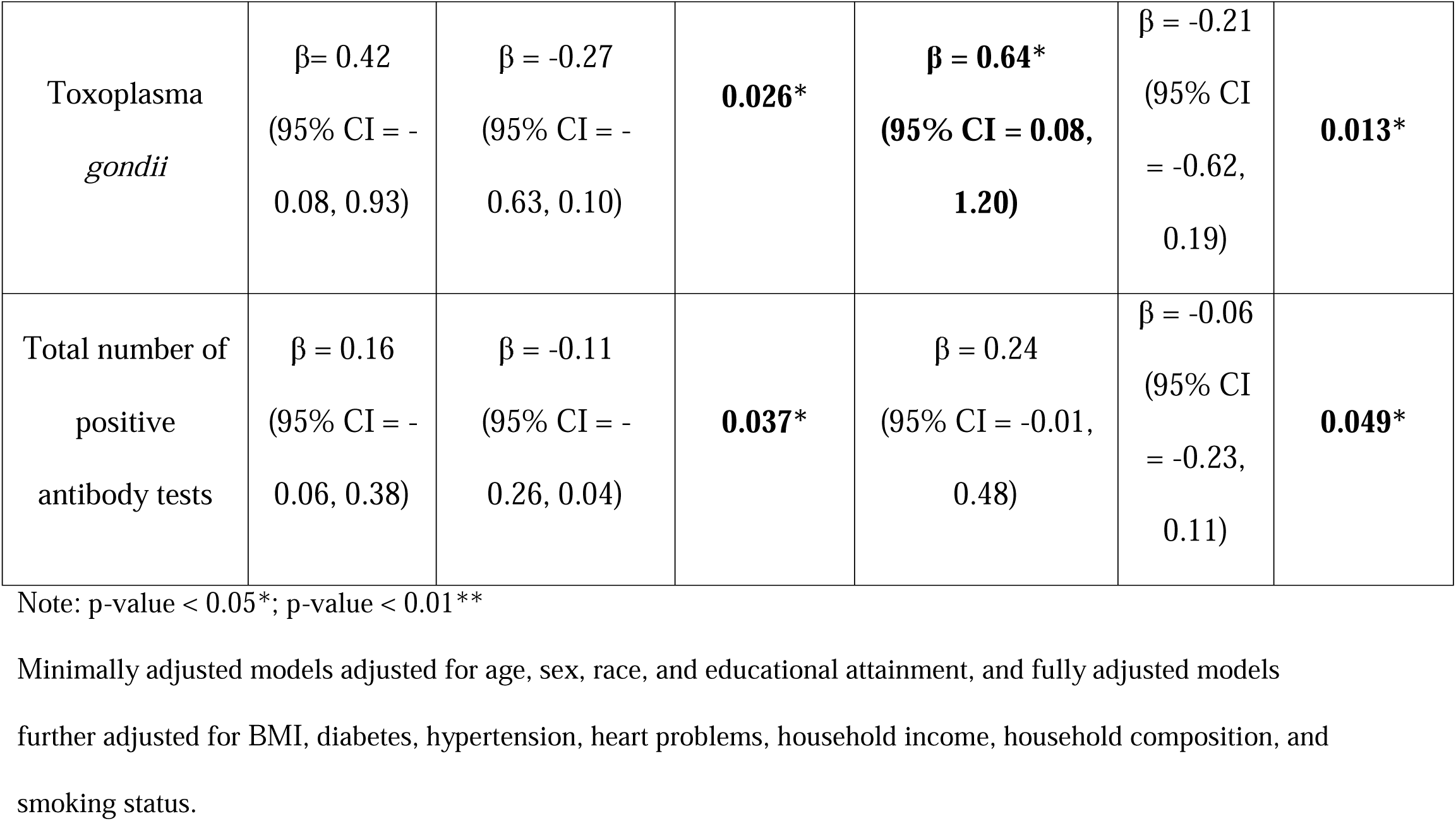
Statistically Significant Interactions of Common Infections with Race in Association with Sleep Outcomes.

|  | Minimally adjusted model |  |  | Fully adjusted model |  |  |
| --- | --- | --- | --- | --- | --- | --- |
|  | Minoritized | White | Interaction<br>p-value | Minoritized | White | Interaction<br>p-value |
| **Insomnia**<br>**symptoms** |  |  |  |  |  |  |
| Cytomegalovirus | **OR =**<br>**0.27\*\***<br>**(95% CI =**<br>**0.11, 0.64)** | OR = 1.49<br>(95% CI = 0.91,<br>2.43) | **0.001\*\*** | **OR = 0.25\*\***<br>**(95% CI = 0.09,**<br>**0.67)** | OR =<br>1.51<br>(95% CI<br>= 0.88,<br>2.60) | **0.002\*\*** |
| **Early awakening** |  |  |  |  |  |  |
| Cytomegalovirus | **OR = 0.39\***<br>**(95% CI =**<br>**0.16, 0.96)** | OR = 1.53<br>(95% CI = 0.86,<br>2.73) | **0.011\*** | **OR = 0.27\***<br>**(95% CI = 0.10,**<br>**0.74)** | OR =<br>1.26<br>(95% CI<br>= 0.67,<br>2.39) | **0.010\*** |
| **Continuous**<br>**sleep duration** |  |  |  |  |  |  |
| Toxoplasma<br>*gondii* | $\beta = 0.42$<br>(95% CI = -<br>0.08, 0.93) | $\beta = -0.27$<br>(95% CI = -<br>0.63, 0.10) | **0.026\*** | **$\beta = 0.64^*$**<br>(95% CI = **0.08,**<br>**1.20)** | $\beta = -0.21$<br>(95% CI<br>= -0.62,<br>0.19) | **0.013\*** |
| Total number of<br>positive<br>antibody tests | $\beta = 0.16$<br>(95% CI = -<br>0.06, 0.38) | $\beta = -0.11$<br>(95% CI = -<br>0.26, 0.04) | **0.037\*** | $\beta = 0.24$<br>(95% CI = -0.01,<br>0.48) | $\beta = -0.06$<br>(95% CI<br>= -0.23,<br>0.11) | **0.049\*** |

Note: p-value < 0.05\*; p-value < 0.01\*\*
Minimally adjusted models adjusted for age, sex, race, and educational attainment, and fully adjusted models
further adjusted for BMI, diabetes, hypertension, heart problems, household income, household composition, and smoking status.

TOX serostatus in relation to sleep duration also varied by race (interaction p-value = 0.013). Among minoritized participants, TOX seropositivity was significantly associated with longer sleep duration (β = 0.64, 95% CI: 0.08, 1.20, p-value = 0.025); Among White participants, TOX was associated with shorter sleep, although this estimate did not reach significance (β = -0.21, 95% CI: -0.62, 0.19, p-value = 0.302). Finally, there was an interaction between race and total number of positive antibody tests (interaction p-value = 0.049). Although point estimates were not significant, they suggest that a greater number of positive antibody tests was associated with a longer sleep duration among minoritized participants (β = 0.24, 95% CI: -0.01, 0.48, p-value = 0.061) but with shorter sleep among White participants (β= -0.06, 95% CI: -0.23, 0.11, p-value = 0.506).

## Discussion

While numerous studies have examined the association of common infections with psychiatric and physical health outcomes,^18,19^ few have examined links of common infections with sleep health, particularly among middle-aged and older adults. In the present study, we found no associations of individual common infections (i.e., CMV, EBV, HSV-1, TOX, VZV) or total number of infections with insomnia, hypersomnia, early awakening, or sleep duration in middle-aged and older adults when White and minoritized participants’ data were examined together. This conflicts with limited research showing that adults 18-80 years of age (M age = 43.63 years, *SD* = 12.56) with insomnia had greater seropositivity for TOX compared to adults without insomnia.^23^ However, our results are consistent with research indicating no differences in insomnia symptoms based on CMV, HSV-1, or TOX seropositivity, and no differences in sleep duration as a function of TOX serostatus among middle-aged and older adults.^33,34^ Discordance in findings could be due to differences in study design, sample composition (e.g., clinical vs. community-based sample), or variability in individual or environmental characteristics (e.g., age, socioeconomic differences, infecting strains of *Toxoplasma*) that may influence associations between infection and sleep health.^50^

We observed numerous interactions between race and common infections with regard to sleep disturbances. CMV seropositivity was associated with 75% lower odds of insomnia symptoms and a 73% lower odds of early awakening among minoritized adults, but associations were in the opposite direction among White adults. In addition, seropositivity for TOX and a greater total infection burden was associated with longer sleep among minoritized participants, but shorter sleep among White participants. Also, infections (i.e. CMV, EBV, total number of positive antibody tests) are more common in minoritized populations in our sample. As noted previously, no prior studies have investigated differences in associations of common infections with sleep by race.

Our results may be attributable to group differences in the tendency to report sleep problems. Participants from minoritized populations have been shown to report fewer sleep complaints than White participants despite potentially having worse sleep when measured objectively.^51^ Because our outcomes are self-reported sleep symptoms, group differences in factors such as responses to stressors, norms surrounding somatic complaints, and stigma may bias infection-sleep associations even if the true association is in fact similar in direction and magnitude. It is also likely that the participants in our study who tested positive for these infections represent a particularly disenfranchised group as most reported a lower household income (shown in the supplementary table 1). There is also evidence that marginalized populations may complain less about sleep problems.^51^ The descriptive statistics in our study also support the differential reporting habits by racial groups (Table 1). Despite a higher burden of health risk factors (e.g., hypertension, lower income), minoritized participants in our sample reported significantly fewer insomnia symptoms (19.2%) than White participants (28.6%). This suggests that the act of reporting a sleep complaint is mediated. Therefore, the lower odds of insomnia among seropositive minoritized individuals may be amplified by this tendency to under-report sleep difficulties or complaints, particularly in a subgroup that may face greater socioeconomic challenges. Objective sleep measures and healthcare professionals verified diagnosis of sleep problems such as insomnia are needed to further adjudicate these patterns.

Another potential explanation for racial/ethnic differences in infection-sleep links may due to the variation in social and environmental exposures, including persistent racism,^52^ historical or contemporary discriminatory housing practices, residential segregation, stress, and socioeconomic deprivation. These factors may contribute to chronic, low-grade inflammation and impairments in immune system functioning,^53^ which may lead to immune adaptation in minoritized participants and make minoritized populations less susceptible to infection-induced sleep disturbances in long term. Over time, minoritized populations may develop a form of immune adaptation or tolerance, whereby the impact of common infections is attenuated. Therefore, CMV seropositivity may have less influence on sleep symptoms in minoritized groups because their immune systems are already conditioned to chronic inflammatory and infectious exposures, whereas in White individuals who are less frequently exposed to these social and environmental exposures, CMV seropositivity may represent a stronger immune challenge that is more likely to manifest as sleep complaints or disrupt sleep.

Our descriptive data further suggest that infection-sleep associations could reflect socioeconomic and health disparities. A greater percentage of CMV seropositive participants in our sample were older, female, and had lower income and education than CMV seronegative individuals, suggesting that CMV serostatus partly reflects cumulative social disadvantage. The majority of minoritized participants were CMV seropositive, with only 16.4% testing negative. Consequently, the protective associations of CMV+ with insomnia and early awakening in minoritized groups may partly reflect selection and reporting bias.

In addition, compared to minoritized populations, White populations may be more sensitive to ambient environmental features such as higher levels of light at night, noise, heat, and air pollution, which can suppress immune system functioning^54–56^ and negatively affect sleep, and perhaps make White individuals more vulnerable to the negative effects of common infection on sleep. Also, White populations may reside in more socioeconomically advantaged areas, have greater health care access and utilization that may modify effects of infections on sleep, overshadowing the detection of significant infection-sleep associations.^57,58^ Future research is needed on potential mechanisms and pathways that may explain racial differences in links between common infections and sleep.

Among minoritized participants, TOX seropositivity and a greater number of positive antibody tests were significantly linked to longer sleep duration, while among White participants those were associated with shorter sleep. The association between a higher infectious burden or TOX seropositivity and longer sleep duration in minoritized participants may reflect poorer overall health. Longer sleep duration is not always good but is instead associated with morbidity, fatigue, and inflammation.^32,33^ Additionally, minoritized participants are more likely to have more comorbidities and worse indicators of health in our sample and also in general.^35^ It is likely that higher seropositivity among minoritized participants reflects a subgroup for whom longer time in bed is driven by fatigue or subclinical illness, rather than by restorative sleep. In contrast, among White participants, the association with shorter sleep may represent a more direct inflammatory response to infection that disrupts sleep. In addition, reporting sleep duration that falls into the long category may not register as a sleep complaint that could be more affected by social perceptions. For sleep duration, one would report hours habitually slept in a more matter of a fact manner than admitting difficulty falling or maintaining sleep. On the other side, prior TOX infection and more common infection-related inflammation or antibodies may contribute to fatigue which will be correlated with longer sleep among minoritized group. Our data indicates that minoritized adults experience a higher burden of hypertension and obesity, along with greater seropositivity for TOX and multiple infections (Table 1). This pattern is consistent with our finding that greater infection burden is associated with longer sleep in minoritized group, as longer sleep maybe a response more suggestive of fatigue and sickness behavior than of restorative sleep. In contrast, White participants exhibit lower overall infection prevalence and cardiometabolic burden (Table 1), seropositivity for CMV or TOX may represent a distinct immune challenge that manifests as shorter sleep in White participants.

Our results have several implications for research and clinical care. That common infections were associated with better and longer sleep among minoritized participants but greater sleep disturbances and shorter sleep among White participants suggests that tailored attention to the presence of common infections may be needed as a means of promoting better overall health across diverse populations. Furthermore, interventions should consider targeting sleep disturbance given the importance of sleep to long-term health and well-being.^59^ Given that overlapping environmental factors likely promote infection exposure and sleep, place-based interventions, such as those that reduce environmental noise, heat, and ambient light, and improve access to green space in urban contexts,^59^ may help offset potential adverse effects of infections on sleep in some White populations, and improve sleep in these communities, independently of infections. Although insomnia symptoms were less reported by minoritized participants in our sample, empirically-supported treatments for insomnia (e.g., cognitive behavior therapy for insomnia; CBT-I),^60^ may also be used to promote sleep health. Moreover, multi-level interventions, including implementation of policies that reduce structural barriers and promote access to health care services, and of routine screenings for and treatment of common infections in clinical-care settings may help mitigate potential effects of common infections with sleep health.

Our study has several limitations. The study design was cross-sectional, which prevented us from examining potential longitudinal, bi-directional associations between common infections and sleep disturbances. Also, the nature of cross-sectional study may introduce reverse causation bias. To permit this, future research should employ longitudinal designs with adequate follow-up time. Also, sleep was assessed via self-report, which may not accurately reflect sleep duration or quality. Specifically, minoritized groups are less likely to report sleep complaints even when objective sleep tends to be worse,^51^ which can bias the infection-sleep associations. Thus, research incorporating objective sleep measures (e.g., polysomnography, actigraphy) is needed. Additionally, common infections and immune functioning correlate with social and environmental factors to some extent.^53–56^Thus, those socioeconomic factors may bias or modify the associations between different ethnic groups, and may bring residual confounding by unmeasured factors. Further, we examined associations of common infections with sleep health in middle-aged and older adults, but it is unclear whether our findings generalize to earlier developmental periods. Another limitation is that we have limited sample size in certain strata (e.g. only 35 participants in CMV negative minoritized group), which may limit the statistical power, constrain the precision, and potentially amplify effect sizes. Although these data were collected between 2003 and 2006, the serological assays used were well-validated and remain in use today,^47,48^ supporting the interpretability of the findings. Nonetheless, changes in infection prevalence, healthcare access, and sleep behaviors may influence the generalizability of the observed associations to more contemporary populations. Future studies using recent cohorts will be important for determining whether these associations have changed over time. Despite these limitations, our study is the first to examine links of a range of common infections and total infection burden with several sleep outcomes and investigate whether race moderates infection-sleep associations.

Our findings of opposite associations between infections and sleep between White and minoritized adults indicate a potential source of sleep disparities that may have implications for other health outcomes and underscore the importance of investigating social determinants of health that may contribute to these disparities. Future research investigating the role of race-based institutional discriminatory policies or interpersonal experiences on sleep disturbances among minoritized populations is needed, as is research focused on identifying factors associated with better sleep quality among White adults seropositive for common infections. The latter may point to interventions to offset potential harms to sleep health induced by infections in minoritized communities and thereby enhance health equity.

## Supporting information

Table 1. Differences in Key Study Variables by Infection Status

## Author Contributions

**Yiwei Yue:** Conceptualization, Methodology, Software, Validation, Formal analysis, Data Curation, Writing - Original Draft, Writing - Review & Editing **Jill A. Rabinowitz:** Conceptualization, Methodology, Formal analysis, Writing - Original Draft, Writing - Review & Editing, Supervision **Chandra L. Jackson:** Writing - Review & Editing **Yiping Xia:** Conceptualization, Writing - Review & Editing **Idiatou Diallo:** Writing - Review & Editing **Robert Yolken:** Conceptualization, Investigation, Resources, Writing - Review & Editing **William W. Eaton:** Writing - Review & Editing **Brion S. Maher:** Conceptualization, Writing - Review & Editing, Supervision, Project administration, and Funding acquisition **Adam P. Spira:** Conceptualization, Writing - Review & Editing, Supervision, Project administration, and Funding acquisition.

## Data Availability Statement

Drs. Adam Spira and Brion Maher will share data from this study with qualified investigators following the completion of data use agreements required by Johns Hopkins University.

