## Supplementary material for "Links of Common Infections with Sleep in Middle-Aged and Older Adults: Modification by Race in the Baltimore Epidemiologic Catchment Area Study": Table 1. Differences in Key Study Variables by Infection Status

| Variable | CMV –  (N = 193) | CMV +  (N = 409) | EBV –  (N = 123) | EBV +  (N = 479) | HSV -  (N = 188) | HSV +  (N = 414) | TOX -  (N = 454) | TOX +  (N = 148) | VZV -  (N = 479) | VZV +  (N = 122) |
| --- | --- | --- | --- | --- | --- | --- | --- | --- | --- | --- |
| Age in years, M (SD) | 54 (10) | 61 (13) | 58 (13) | 59 (13) | 56 (12) | 60 (13) | 56 (11) | 67 (14) | 59 (13) | 61 (14) |
| Race, N (%) | | | | | | | | | | |
| White | 158 (82%) | 230 (56%) | 98 (80%) | 290 (61%) | 123 (65%) | 265 (64%) | 288 (63%) | 100 (68%) | 311 (65%) | 76 (62%) |
| Minoritized | 35 (18%) | 179 (44%) | 25 (20%) | 189 (39%) | 65 (35%) | 149 (36%) | 166 (37%) | 48 (32%) | 168 (35%) | 46 (38%) |
| Sex, N (%) | | | | | | | | | | |
| Male | 98 (51%) | 124 (30%) | 46 (37%) | 176 (37%) | 84 (45%) | 138 (33%) | 176 (39%) | 46 (31%) | 180 (38%) | 42 (34%) |
| Female | 95 (49%) | 285 (70%) | 77 (63%) | 303 (63%) | 104 (55%) | 276 (67%) | 278 (61%) | 102 (69%) | 299 (62%) | 80 (66%) |
| Household size, M (SD) | 3.12 (1.32) | 3.09 (1.49) | 3.28 (1.52) | 3.05 (1.41) | 3.12 (1.39) | 3.08 (1.46) | 3.09 (1.35) | 3.10 (1.70) | 3.12 (1.45) | 3.03 (1.41) |
| Annual household income, M (SD) | 19.4 (4.1) | 16.5 (5.3) | 18.4 (4.4) | 17.2 (5.3) | 18.4 (4.6) | 17.0 (5.3) | 17.7 (5.1) | 16.6 (5.1) | 17.6 (5.2) | 16.7 (5.0) |
| Grade, M (SD) | 13.08 (2.30) | 12.03 (2.82) | 12.32 (2.53) | 12.38 (2.76) | 12.90 (2.81) | 12.12 (2.63) | 12.66 (2.56) | 11.45 (2.97) | 12.45 (2.67) | 11.99 (2.82) |
| BMI, M (SD) | 30 (7) | 30 (8) | 30 (7) | 30 (8) | 30 (9) | 30 (7) | 30 (8) | 29 (7) | 30 (7) | 30 (9) |
| Diabetes, N (%) | | | | | | | | | | |
| No | 159 (82%) | 338 (83%) | 98 (80%) | 399 (83%) | 167 (89%) | 330 (80%) | 377 (83%) | 120 (81%) | 395 (83%) | 101 (83%) |
| Yes | 34 (18%) | 70 (17%) | 25 (20%) | 79 (17%) | 21 (11%) | 83 (20%) | 76 (17%) | 28 (19%) | 83 (17%) | 21 (17%) |
| Hypertension, N (%) | | | | | | | | | | |
| No | 113 (59%) | 192 (47%) | 65 (53%) | 240 (51%) | 110 (59%) | 195 (48%) | 229 (51%) | 76 (53%) | 241 (51%) | 63 (53%) |
| Yes | 77 (41%) | 213 (53%) | 57 (47%) | 233 (49%) | 77 (41%) | 213 (52%) | 222 (49%) | 68 (47%) | 234 (49%) | 56 (47%) |
| Heart problems, N (%) | | | | | | | | | | |
| No | 159 (83%) | 334 (82%) | 101 (83%) | 392 (83%) | 160 (85%) | 333 (81%) | 379 (84%) | 114 (78%) | 392 (83%) | 100 (82%) |
| Yes | 33 (17%) | 71 (18%) | 21 (17%) | 83 (17%) | 28 (15%) | 76 (19%) | 72 (16%) | 32 (22%) | 82 (17%) | 22 (18%) |
| Smoking status, N (%) | | | | | | | | | | |
| Non-smokers | 117 (61%) | 229 (57%) | 82 (67%) | 264 (56%) | 104 (56%) | 242 (59%) | 258 (57%) | 88 (62%) | 276 (58%) | 70 (58%) |
| Former smokers | 12 (6%) | 49 (12%) | 13 (10%) | 48 (10%) | 18 (10%) | 43 (11%) | 42 (9%) | 19 (13%) | 46 (10%) | 14 (11%) |
| Current smokers | 62 (33%) | 126 (31%) | 28 (23%) | 160 (34%) | 64 (34%) | 124 (30%) | 152 (34%) | 36 (25%) | 151 (32%) | 37 (31%) |
| Insomnia symptoms, N (%) | | | | | | | | | | |
| No | 136 (72%) | 304 (75%) | 83 (67%) | 357 (76%) | 133 (72%) | 307 (75%) | 330 (73%) | 110 (77%) | 354 (75%) | 86 (71%) |
| Yes | 53 (28%) | 99 (25%) | 40 (33%) | 112 (24%) | 51 (28%) | 101 (25%) | 120 (27%) | 32 (23%) | 116 (25%) | 35 (29%) |
| Early awakening, N (%) | | | | | | | | | | |
| No | 159 (83%) | 328 (81%) | 98 (80%) | 389 (82%) | 156 (84%) | 331 (81%) | 368 (81%) | 119 (83%) | 393 (83%) | 93 (77%) |
| Yes | 32 (17%) | 76 (19%) | 25 (20%) | 83 (18%) | 30 (16%) | 78 (19%) | 84 (19%) | 24 (17%) | 80 (17%) | 28 (23%) |
| Hypersomnia symptoms, N (%) | | | | | | | | | | |
| No | 174 (92%) | 379 (94%) | 113 (92%) | 440 (93%) | 176 (95%) | 377 (92%) | 421 (93%) | 132 (93%) | 442 (93%) | 110 (92%) |
| Yes | 16 (8.4%) | 25 (6.2%) | 10 (8.1%) | 31 (6.6%) | 9 (4.9%) | 32 (7.8%) | 31 (6.9%) | 10 (7.0%) | 31 (6.6%) | 10 (8.3%) |
| Total sleep time, M (SD) | 7.25 (6.73) | 7.58 (8.00) | 6.80 (1.41) | 7.65 (8.50) | 7.79 (9.53) | 7.33 (6.56) | 7.22 (6.25) | 8.29 (10.83) | 7.05 (4.46) | 9.14 (14.31) |

Note. CMV = Cytomegalovirus, EBV = Ebstein Barr Virus, HSV = Herpes Simplex Virus-1, TOX = Toxoplasma *gondii*, VZV = Varicella Zoster Virus

**Table 1.** Study Population Characteristics Overall and by Race, Baltimore Epidemiologic Catchment Area (ECA) study Wave 4 (2003-2006)

| **Characteristic** | **Total** | **White** | **Minoritized^1^** | **p-value** |
| --- | --- | --- | --- | --- |
|  | **N=602** | **N=388** | **N=214** |  |
| **Age, Mean (SD), years** | 59.03 (12.80) | 59.75 (13.46) | 57.73 (11.43) | 0.064 |
| **Race, N (%)** |  |  |  |  |
| White adults | 388 (64.5%) | - | - | - |
| Minoritized adults | 214 (35.6%) | - | - | - |
| **Sex, N (%)** |  |  |  | **<0.001** |
| Male | 222 (36.9%) | 165 (42.5%) | 57 (26.6%) |  |
| Female | 380 (63.1%) | 223 (57.5%) | 157 (73.4%) |  |
| **Educational attainment,**  **M (SD)** | 12.37 (2.71) | 12.49 (2.63) | 12.14 (2.84) | 0.12 |
| **Body Mass Index, M (SD)** | 30.01 (7.68) | 29.19 (6.68) | 31.44 (9.02) | **0.001** |
| **Diabetes, N (%)** |  |  |  | 0.80 |
| No | 497 (82.6%) | 322 (83.0%) | 175 (81.8%) |  |
| Yes | 104 (17.3%) | 66 (17.0%) | 38 (17.8%) |  |
| Missing | 1 (0.2%) | 0 (0.0%) | 1 (0.5%) |  |
| **Hypertension, N (%)** |  |  |  | **<0.001** |
| No | 305 (50.7%) | 217 (55.9%) | 88 (41.1%) |  |
| Yes | 290 (48.2%) | 167 (43.0%) | 123 (57.5%) |  |
| Missing | 7 (1.2%) | 4 (1.0%) | 3 (1.4%) |  |
| **Heart problems, N (%)** |  |  |  | **0.012** |
| No | 493 (81.9%) | 306 (78.9%) | 187 (87.4%) |  |
| Yes | 104 (17.3%) | 78 (20.1%) | 26 (12.1%) |  |
| Missing | 5 (0.8%) | 4 (1.0%) | 1 (0.5%) |  |
| **Annual household income,**^2^ **M (SD)** | 17.45 (5.15) | 18.48 (4.53) | 15.59 (5.68) | **<0.001** |
| **Household size, M (SD)** | 3.10 (1.44) | 3.02 (1.40) | 3.23 (1.50) | 0.081 |
| **Insomnia symptoms, N (%)** |  |  |  | **0.011** |
| No | 440 (73.1%) | 271 (69.8%) | 169 (79.0%) |  |
| Yes | 152 (25.2%) | 111 (28.6%) | 41 (19.2%) |  |
| Missing | 10 (1.7%) | 6 (1.5%) | 4 (1.9%) |  |
| **Early awakening, N (%)** |  |  |  | 0.74 |
| No | 487 (80.9%) | 312 (80.4%) | 175 (81.8%) |  |
| Yes | 108 (17.9%) | 71 (18.3%) | 37 (17.3%) |  |
| Missing | 7 (1.2%) | 5 (1.3%) | 2 (0.9%) |  |
| **Hypersomnia symptoms, N (%)** |  |  |  | 0.83 |
| No | 553 (91.9%) | 355 (91.5%) | 198 (92.5%) |  |
| Yes | 41 (6.8%) | 27 (7.0%) | 14 (6.5%) |  |
| Missing | 8 (1.3%) | 6 (1.5%) | 2 (0.9%) |  |
| **Total sleep time, M (SD)** | 6.86 (1.53) | 6.86 (1.43) | 6.87 (1.69) | 0.95 |
| **Herpes simplex virus-1, N (%)** |  |  |  | 0.74 |
| Negative | 188 (31.2%) | 123 (31.7%) | 65 (30.4%) |  |
| Positive | 414 (68.8%) | 265 (68.3%) | 149 (69.6%) |  |
| **Cytomegalovirus, N (%)** |  |  |  | **<0.001** |
| Negative | 193 (32.1%) | 158 (40.7%) | 35 (16.4%) |  |
| Positive | 409 (67.9%) | 230 (59.3%) | 179 (83.6%) |  |
| **Epstein-Barr virus, N (%)** |  |  |  | **<0.001** |
| Negative | 123 (20.4%) | 98 (25.3%) | 25 (11.7%) |  |
| Positive | 479 (79.6%) | 290 (74.7%) | 189 (88.3%) |  |
| **Varicella zoster virus, N (%)** |  |  |  | 0.59 |
| Negative | 122 (20.3%) | 76 (19.6%) | 46 (21.5%) |  |
| Positive | 479 (79.6%) | 311 (80.2%) | 168 (78.5%) |  |
| Missing | 1 (0.2%) | 1 (0.3%) | 0 (0.0%) |  |
| **Toxoplasma *gondii*, N (%)** |  |  |  | 0.36 |
| Negative | 454 (75.4%) | 288 (74.2%) | 166 (77.6%) |  |
| Positive | 148 (24.6%) | 100 (25.8%) | 48 (22.4%) |  |
| **Total number of positive antibody tests, M (SD) (range)** | 2.62 (1.07) (range: 0-5) | 2.48 (1.10) (range: 0-5) | 2.86 (0.96) (range: 0-5) | **<0.001** |

^1^Minoritized adults included individuals who identified as either American Indian, Alaskan Native, Asian, Pacific Islander, Black not Hispanic, Hispanic, or White not Hispanic.

^2^Based on the income categories, 17.45 reflects an average income ranging from $25,000-$34,999 for the whole sample; 18.48 reflects an average income ranging from $35,000-49,999 among White adults; and 15.59 reflects an average income ranging $17,500-$19,999.

**Table 2.** Associations between Common Infections and Sleep Disturbances, β (95% CI)

|  | **Minimally adjusted model** | **Fully adjusted model** |
| --- | --- | --- |
| **Insomnia symptoms** |  | |
| Cytomegalovirus | 1.05 (0.67, 1.63) | 1.05 (0.64, 1.72) |
| Epstein-Barr virus | 0.73 (0.47, 1.14) | 0.66 (0.40, 1.10) |
| Herpes simplex virus-1 | 0.88 (0.58, 1.32) | 0.77 (0.49, 1.21) |
| Varicella zoster virus | 1.34 (0.85, 2.12) | 1.52 (0.91, 2.54) |
| Toxoplasma *gondii* | 0.93 (0.57, 1.51) | 1.10 (0.64, 1.88) |
| Total number of positive antibody tests | 0.97 (0.79, 1.19) | 0.98 (0.78, 1.22) |
| **Early awakening** |  | |
| Cytomegalovirus | 1.09 (0.66, 1.81) | 0.87 (0.49, 1.52) |
| Epstein-Barr virus | 0.86 (0.52, 1.43) | 0.78 (0.44, 1.39) |
| Herpes simplex virus-1 | 1.15 (0.72, 1.85) | 0.96 (0.57, 1.61) |
| Varicella zoster virus | 1.45 (0.89, 2.37) | 1.54 (0.89, 2.69) |
| Toxoplasma *gondii* | 0.80 (0.47, 1.37) | 0.90 (0.49, 1.64) |
| Total number of positive antibody tests | 1.05 (0.84, 1.31) | 0.98 (0.76, 1.26) |
| **Hypersomnia** |  | |
| Cytomegalovirus | 0.76 (0.36, 1.60) | 0.57 (0.24, 1.35) |
| Epstein-Barr virus | 0.83 (0.39, 1.79) | 0.71 (0.28, 1.78) |
| Herpes simplex virus-1 | 1.76 (0.80, 3.83) | 2.53 (0.98, 6.50) |
| Varicella zoster virus | 1.37 (0.64, 2.91) | 1.26 (0.52, 3.06) |
| Toxoplasma *gondii* | 1.37 (0.62, 3.05) | 1.89 (0.74, 4.78) |
| Total number of positive antibody tests | 1.15 (0.82, 1.61) | 1.18 (0.78, 1.79) |
| **Sleep duration** |  | |
| Cytomegalovirus | -0.08 (-0.37, 0.21) | -0.05 (-0.36, 0.27) |
| Epstein-Barr virus | 0.07 (-0.24, 0.37) | 0.13 (-0.22, 0.47) |
| Herpes simplex virus-1 | -0.03 (-0.30, 0.24) | 0.06 (-0.23, 0.35) |
| Varicella zoster virus | -0.05 (-0.36, 0.26) | -0.02 (-0.36, 0.32) |
| Toxoplasma *gondii* | -0.04 (-0.34, 0.27) | 0.08 (-0.26, 0.42) |
| Total number of positive antibody tests | -0.03 (-0.16, 0.10) | 0.03 (-0.11, 0.18) |

Note: p-value < 0.05*; p-value < 0.01**

Minimally adjusted models adjusted for age, sex, race, and educational attainment, and fully adjusted models further adjusted for BMI, diabetes, hypertension, heart problem, household income, household composition, and smoking status.

**Table 3.** Statistically Significant Interactions of Common Infections with Race in Association with Sleep Outcomes

|  | **Minimally adjusted model** | | | **Fully adjusted model** | | |
| --- | --- | --- | --- | --- | --- | --- |
|  | **Minoritized** | **White** | **Interaction p-value** | **Minoritized** | **White** | **Interaction p-value** |
| **Insomnia symptoms** |  | | | | | |
| Cytomegalovirus | **OR = 0.27** (95% CI = 0.11, 0.64)** | OR = 1.49  (95% CI = 0.91, 2.43) | **0.001**** | **OR = 0.25** (95% CI = 0.09, 0.67)** | OR = 1.51  (95% CI = 0.88, 2.60) | **0.002**** |
| **Early awakening** |  |  |  |  |  |  |
| Cytomegalovirus | **OR = 0.39***  **(95% CI = 0.16, 0.96)** | OR = 1.53  (95% CI = 0.86, 2.73) | **0.011*** | **OR = 0.27***  **(95% CI = 0.10, 0.74)** | OR = 1.26  (95% CI = 0.67, 2.39) | **0.010*** |
| **Continuous sleep duration** |  | | | | | |
| Toxoplasma *gondii* | β= 0.42  (95% CI = -0.08, 0.93) | β = -0.27  (95% CI = -0.63, 0.10) | **0.026*** | **β = 0.64***  **(95% CI = 0.08, 1.20)** | β = -0.21  (95% CI = -0.62, 0.19) | **0.013*** |
| Total number of positive antibody tests | β = 0.16  (95% CI = -0.06, 0.38) | β = -0.11  (95% CI = -0.26, 0.04) | **0.037*** | β = 0.24  (95% CI = -0.01, 0.48) | β = -0.06  (95% CI = -0.23, 0.11) | **0.049*** |

Note: p-value < 0.05*; p-value < 0.01**

Minimally adjusted models adjusted for age, sex, race, and educational attainment, and fully adjusted models further adjusted for BMI, diabetes, hypertension, heart problems, household income, household composition, and smoking status.
